# Geographic barriers to establishing a successful hospital referral system in rural Madagascar

**DOI:** 10.1101/2021.08.06.21261682

**Authors:** Felana Angella Ihantamalala, Matthew H Bonds, Mauricianot Randriamihaja, Luc Rakotonirina, Vincent Herbreteau, Christophe Révillion, Serge Rakotoarimanana, Giovanna Cowley, Tsirinomen’ny Aina Andritiana, Alishya Mayfield, Michael Rich, Rado JL Rakotonanahary, Karen E Finnegan, Andriamanolohaja Ramarson, Benedicte Razafinjato, Bruno Ramiandrisoa, Andriamihaja Randrianambinina, Laura F Cordier, Andres Garchitorena

**Affiliations:** NGO PIVOT, Ranomafana, Madagascar; Department of Global Health and Social Medicine, Harvard Medical School, Boston, USA; Espace-Dev, IRD, Univ Antilles, Univ Guyane, Univ Montpellier, Univ Réunion, Phnom Penh, Cambodia; Espace-Dev, IRD, Univ Antilles, Univ Guyane, Univ Montpellier, Univ Réunion, Saint-Pierre, La Réunion, France; Division of Global Health Equity, Brigham and Women’s Hospital, USA; Partners In Health, USA; Ministry of Health in Madagascar, Antananarivo, Madagascar; MIVEGEC, Univ. Montpellier, CNRS, IRD, Montpellier, France

## Abstract

**Background:** The provision of emergency and hospital care has become an integral part of the global vision for universal health coverage. It is recommended that at least 80% of a country’s population should be within two hours of a facility with essential surgery and anesthetic services. In order to strengthen health systems to achieve this goal, there needs to be an understanding of the time necessary for populations to reach a hospital. The goal of this study was to develop methods that accurately estimate referral and pre-hospital time for rural health districts in LMICs. We used these estimates to assess how the local geography can limit the impact of a strengthened referral program in a rural district of Madagascar.

**Methods:** We developed a database containing 1) travel speed in a subset of routes by foot and motorized vehicles in Ifanadiana District; 2) a full mapping of all roads, footpaths and households in the district; and 3) remotely sensed data on terrain, land cover and climatic characteristics. We used this information to calibrate estimates of referral and pre-hospital time based on shortest route algorithms and statistical models of local travel speed. We compared these estimates with those from other commonly used methods in geographic accessibility modeling. Finally, we studied the impact of referral time on the evolution of the number of referrals completed by each health center in the district in 2014-2020 via generalized linear mixed models, using model estimates to predict the impact on referral numbers of strategies aimed at reducing referral time for underserved populations.

**Results:** About 10% of the population lived less than two hours from the hospital, and more than half lived over four hours away, with variable access depending on climatic conditions. Only the four health centers (out of 21) located near the paved road had referral times to the hospital within one hour, which contributed over 75% of all 8,464 hospital referrals. Referral time remained the main barrier limiting the number of referrals despite health system strengthening efforts. The addition of two new referral centers is estimated to triple the population living within two hours from a center with higher acute care capacity and nearly double the number of referrals expected.

**Conclusion:** This study demonstrates how adapting geographic accessibility modeling methods to local scales can occur through improving the precision of travel time estimates and pairing them with data on health facility data. Such information can substantially improve the design of a local health system to overcome existing barriers to care and achieve universal health coverage.

## Introduction

The provision of emergency care has recently become an integral part of the global vision for Universal Health Coverage (UHC), as a way to ensure the “timely and effective delivery of life-saving health care services to those in need”.(1) While primary care services remain the cornerstone of any health system, it is estimated that nearly half of all deaths and about a third of disability-adjusted life years in low and middle income countries (LMICs) could be addressed by well-functioning emergency care systems.(2) The 2019 World Health Assembly urged Member States to strengthen emergency care systems by creating policies, governance, and funding mechanisms that promote the inclusion of emergency care into national health strategies.(1) These systems should overcome the multiple geographical, financial and social barriers that limit the access of patients in LMICs to hospital care.(3–7) In rural areas specifically, where healthcare infrastructure is sparse, geographic access to care is a key determinant of hospital use, which tends tofall exponentially as the travel time to health facilities increases.(8–10) To improve geographic access to hospital care, the Lancet Commission on Global Surgery proposed that at least 80% of a country’s population should be within two hours of a facility with essential surgery and anesthetic services.(11)

A variety of methods exist for estimating populations’ time to reach the nearest hospital; these tend to differ according to the study setting. In low income countries, where road infrastructure and emergency services are well developed, travel time is typically estimated using shortest route estimations, with assumptions about vehicle travel speeds.(12–15) To improve on these methods, estimates are often compared or calibrated using surveys of referred patients,(12–14) trip data from emergency service records,(13) or ambulance global positioning system (GPS)measurements.(16,17) In contrast, studies in rural areas of low-income countries combine estimates of travel time by foot and motorized vehicles to reflect local realities in hospital accessibility, as road infrastructure is deficient and ambulance services are rarely available.

Since much of the geographic information on roads and paths that people use to reach hospitals in these areas is still missing, the prevailing technique for estimating travel time is the use of friction surfaces based on terrain and land cover characteristics, together with available road networks.(8,18–21) For instance, these algorithms are the basis of “AccessMod”, a free tool developed by the World Health Organization (WHO) to increase the adoption of geographic accessibility analyses into health planning. (22,23) Using such methods, researchers have estimated that over 70%(24) and as much as 92%(25) of the population in sub-Saharan Africa live within two hours of a hospital. However, methods that rely on friction surfaces represent only a “best guess” of the routes people use, with simplifying assumptions that rarely account for the impact of context-specific factors on travel speeds or on accessibility at the local level. As a result, current estimates of hospital accessibility in sub-Saharan Africa could be grossly overestimated.

To help increase access to timely emergency and hospital care in LMICs, health system strengthening efforts need to integrate emergency care training, processes, and resources at different levels of the health system.(1,11) The WHO recommends that, patients be transferred from primary health centers to referral hospitals by ambulances in emergency cases.(26) In practice, formal emergency care systems in LMICs are rare and poorly implemented due to the lack of resources (skilled personnel, infrastructure, transport) to ensure timely provision to emergency services.(27) Even where ambulances are available, time-delays in hospital transfers limits the coverage and effectiveness of emergency services,(4,8) increasing the risk of death for children,(28) pregnant women with delivery complications,(29) or patients with conditions such as stroke(30) or asthma.(31) Given the importance of ensuring timely access to emergency and hospital care, accurate estimations of referral and pre-hospital time are key, not just to identify global needs and funding gaps, but also to improve the basic design of health systems at the national and local levels in LMICs.

The goal of this study was to develop methods that accurately estimate referral and pre-hospital time for rural health districts in LMICs, based on locally calibrated models and context-specific information. We used these estimates to assess how local geography can limit the impact of a strengthened referral program and which additional strategies can help overcome geographic barriers. This study was conducted in Ifanadiana District, in southeastern Madagascar, which has been established as a model district for UHC by the government of Madagascar and the healthcare no-governmental organization (NGO) PIVOT (see “Study area and intervention”). Ifanadiana District has one of the only rural ambulance referral systems in the country. We build on previous work in which we improved the precision of geographic accessibility estimates to primary care, showing that standard methods tend to overestimate the proportion of the population with access to care.(32) We hypothesized that estimates of hospital access might be subject to a similar or greater overestimation, and that even small differences in time to access hospital care can have substantial impacts on the use of emergency services.

To address this, we first developed a database containing the movements of people and motorized vehicles in Ifanadiana District over several months as well as a full mapping of all roads, footpaths and households in the district. We used this information to calibrate estimates of referral and pre-hospital time based on shortest route algorithms and statistical models of local travel speed. Finally, we studied the impact of referral time on the evolution of the number of referrals completed by each health center in the district, using model estimates to predict the impact on referral numbers of strategies aimed at reducing referral time for underserved populations.

## Methods

### Study area and intervention

Ifanadiana is a rural district located in Southeastern Madagascar. Ifanadiana has 15 communes and 195 fokontany (smallest administrative boundary comprising one or several villages). The district health system includes one referral hospital and 21 health centers, including a primary care health center (CSB2) in each commune, and 6 additional basic health centers (CSB1) in some of the larger communes, which provide a more limited range of primary care services. In addition, two community health workers per fokontany provide care for common illnesses of children under 5 years. The MoPH-PIVOT model district is based on strengthening the health system at all three levels of care, ensuring i) health system readiness via the availability of sufficient health personnel, essential medicines, necessary equipment, and infrastructure renovations; ii) the provision of high-quality accessible care, including improved diagnostic capacity, clinical programs, infection control, psychosocial assistance, the removal of user-fees, and the distribution of social kits to the patients; and iii) integrated data systems at all levels of care to allow for robust monitoring, evaluation and research on the intervention. More information about the health system strengthening intervention can be found in the supplementary table S1.

To improve access to emergency and hospital care, the MoPH-PIVOT partnership created an ambulance referral system, which operates 24 hours a day and 7 days a week. This service includes three fully equipped ambulances with 12 trained paramedics, and processes to ensure care continuity. For instance, all health centers in the district have been equipped with cell phones and a call center has been established at the district hospital for specialized guidance and to coordinate referrals.(33) Patients requiring secondary care are transported from accessible health centers or meeting points to the district hospital, and those who need more specialized care are transported to the university hospital in the city of Fianarantsoa (two hours away by car) or to specialized facilities in Antananarivo (10 hours away by car). In non-emergency cases needing secondary care, other means of transport can be used by patients (such as local bus: “*taxi brousse*”) and transportation fees are reimbursed by PIVOT.

Multiple geographic barriers limit the success of this referral program. The district is made up of deep valleys and a mountainous landscape, with an altitude gradient ranging from 1000 m to 100 m from West to East. Road infrastructure in the district is scarce (figure 1), with a single paved road that runs across the district from East to West through four of its communes. In addition, there are two main non-paved roads connecting the district capital, Ifanadiana, with the communes in the North and South. Only a fraction of these roads is accessible by all-terrain vehicles, especially under rainy conditions, and the availability of such vehicles is very limited. The rest of the road network consists of smaller paths accessible by all-terrain motorcycles, with most of the network accessible only by foot. To further improve access to emergency and secondary care, the MoPH-PIVOT partnership is planning to turn two major health centers in the North and South of the district into referral centers that will be able to provide acute care for patients in distress. This research was done in support of this referral program, to accurately estimate current accessibility to secondary care and to assess the potential impact of this strategy at addressing existing challenges.

**Figure 1.**
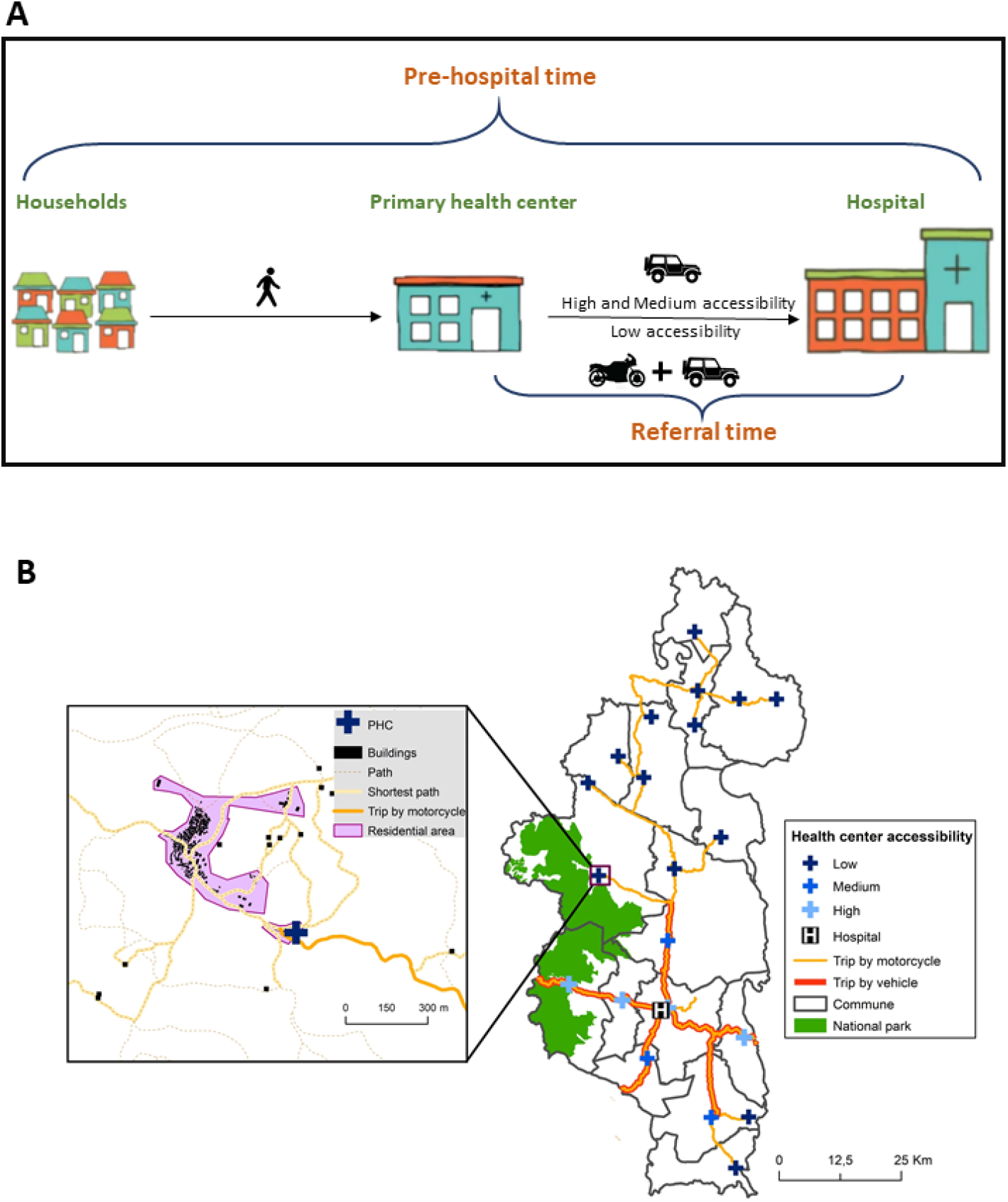
Pre-hospital circuit in Ifanadiana. A) Shows the different routes of patients from households to the hospital in case of referral B) The main road to join primary health centers (PHCs) and the district hospital.

### Data collection

#### Road and footpath network in Ifanadiana District

To accurately estimate pre-hospital time from each household in Ifanadiana District to the referral hospital, we first used the OpenStreetMap (OSM)(34) platform to map the full network of roads and footpaths, as well as all residential areas, houses and rice fields in the district. This was done in collaboration with the Humanitarian OpenStreetMap Team (HOT),(35) which allowed us to divide the district into several thousand 1km^2^ tiles that were then mapped by individual users using very high-resolution satellite images. The mapping of each tile was done in two steps: first, a user mapped all geographic entities and then another user validated the mapping to ensure data quality. As a result, over 20,000 km of footpaths and over 100,000 buildings were mapped. The purpose of obtaining this dataset was to determine the shortest path between each household and its nearest health center, and between each health center and the district hospital, which emulates the process of a referral from the community to the hospital (figure 1). Distance estimations were done via the Open-Source Routing Machine (OSRM) tool. This work is extensively described in Ihantamalala *et al*. 2020.

#### Travel speed under different terrains and conditions for people and motorized vehicles

To obtain context-specific estimates of travel speed under different local terrain and climatic conditions, we collected two types of datasets. First, to estimate travel speeds by foot, from September 2018 to April 2019, we recorded the movements of different individuals (NGO staff and community health workers) in a representative sample of footpaths in Ifanadiana District. For this, we used the android version 3.0.2 of the OSMAnd app(36) installed in Samsung Tab A10 tablets, using a record frequency of 10 seconds. For each GPS point the following data was recorded: geographical location, date, time and altitude. Overall, 168 different routes were recorded, representing nearly 1,000 km of footpaths. This dataset was used to estimate the time it takes for patients to reach the nearest health center prior to referral, and is described in further detail in Ihantamalala *et al*. 2020.

Second, to estimate travel speeds for motorized vehicles (cars and motorcycles), we obtained trip recordings of all the NGO cars and motorcycles from TAG-IP,(37) a company that provides vehicle GPS data collection and management services in Madagascar. All the NGO motorized vehicles are equipped with one of three types of GPS: Careu U1, Careu Uw1 or FMB204. These GPS devices were calibrated to record location every 10 seconds and data were transferred and stocked directly into the server of TAG-IP. For this project, we obtained a sample of data from March 17 to May 5, 2019 and from February 1 to April 4, 2020. A total of 77,709 km of motorized vehicle routes from Ifanadiana District were collected, 61,258 km from cars and 16,451 km from motorcycles (supplementary table S2). For each GPS point, the following data were recorded: identity of the car/motorcycle coordinates of their position, date, time, and altitude. This dataset was then used to estimate the referral time of patients from each health center to the district hospital.

#### Satellite imagery and remote sensing

We obtained elevation and precipitation data from remotely sensed data. We downloaded the Shuttle Radar Topography Mission (SRTM) Digital Elevation Model (DEM) from the United States Geological Survey (USGS,(38)), which gives elevation with a 30 m ground resolution. We also acquired precipitation estimates from the NASA Prediction of Worldwide Energy Resources (POWER) Project,(39) with a spatial resolution of 0.5 * 0.5 degrees.

#### Referral data

The MoPH-PIVOT partnership collected hospital data on the number of patients referred from health centers to the hospital, with data accessible via the NGO’s dashboard(40) for use by program managers. We obtained data from PIVOT’s monitoring and evaluation team for all referrals completed from January 2014 to July 2020. The data included the aggregated number of referrals per month, means of transportation used, and the name of the health center from which the referral originated. A referral was considered complete when the patient reached the hospital and feedback was provided to the referring health center. Personal identifiable information from all referral data was removed to protect patient privacy. Use of health system data for this study was approved by the Medical Inspector of Ifanadiana District, and by Harvard’s Institutional Review Board.

### Data analysis

#### Estimation of referral time from health centers and households to the district hospital

This work builds on previous work where we estimated the travel time from every household in Ifanadiana District to the nearest health center.(32) Here we estimate total pre-hospital time for a referral by adding the time from each health center to the district hospital. To calculate the referral time between health centers and the hospital, we used the database of the NGO’s vehicle movements and divided each trip into 100 m segments which we intersected with geographic and climatic data. We explored the impact on travel speed of: the type of motorized vehicle, the type of road, rainfall conditions (one-month average of precipitation, in mm), driving through residential areas, crossing bridges, and the distance since the trip began (supplementary table S3). Initially, we carried out exploratory and univariate analyses to understand the relationship between travel speed and each variable of interest, transforming explanatory variables where necessary. Then, we carried out a multivariate analysis using a Poisson mixed-effects model that included all of the above variables as fixed effects and each separate trip as a random intercept. Based on this model, we predicted referral time for each of the 21 health centers according to the particular route characteristics to reach the hospital, and two types of rainfall conditions: dry and rainy conditions. For the predictions under rainy conditions, we used the median value of rainfall recorded during the vehicle trips.

Second, we combined travel time from health centers to the hospital with the travel time from households to health centers estimated in our previous research.(32) In brief, travel time to the nearest health center was estimated using the same methods as described above, where a statistical model of travel speed by foot was first built from field data as a function of land cover, climatic and elevation characteristics. Then, predictions were made for all of the 41,426 shortest routes from each household or residential area in the district to the nearest health center, under both dry and rainy conditions. Our estimates of pre-hospital time assume that a patient went to the closest health center by foot, and was then transferred via ambulance if the health center was accessible by car, and if not, by other means of transport (in our case by motorcycle) to an ambulance pick-up point (figure 1). Ambulances and cars can drive on the paved road, and 17 km north and 16km south of the paved road on non-paved roads (figure 1). These assumptions are based on the reports by patients and healthcare providers in Ifanadiana.

Finally, we aggregated individual household pre-hospital times to estimate the percentage of the population at different categories of access to the district hospital. We also interpolated the estimates of pre-hospital time between households using kriging methods available on ArcGIS to visualize its spatial distribution over the entire district.

#### Comparison of pre-hospital time results with existing methods

To assess how the locally calibrated estimates obtained in our study compared to other methods commonly used in geographical accessibility modeling of hospital care, we estimated referral and pre-hospital time using simplified assumptions generally accepted for similar settings. For referral time we used the same shortest route, but with a simplified assumption that vehicles travel at a speed of 50 km/h on paved secondary roads and 30km/h on non-paved roads. For estimated of pre-hospital time, we used the software “AccesMod”, which is based on friction surfaces algorithms, together with the district’s digital elevation model, OSM road network and land cover datasets, where speed values for each class of land cover and road network were assigned following recommendations for AccessMod 3.0.(22)

#### Assessment of determinants of monthly referrals by health centers

We assessed the evolution of the number of referrals completed by health centers since the beginning of the referral program to understand the impact of geographic barriers. For this, we first classified health centers into four categories according to their geographic accessibility: high for those on the paved road (N=4), medium for those accessible by ambulance on a non-paved road (N=3), and low for those not accessible by ambulance and requiring additional means of transportation (N=14). The latter were further subdivided into major (CSB2, N=8) or basic health centers (CSB1, N=6). We explored the number of referrals completed and the means of transportation used for each category of geographic accessibility.

Second, we used multivariate statistical models to study how geographic factors affected the monthly number of referrals completed by each health center, adjusting for relevant variables. We assessed the impact of geographic factors via orthogonal polynomial terms of degree 2 (non-linear relationship) for the travel time between the health center and the district hospital, under both rainy and dry conditions, including the categories of access described above. We adjusted the model for health system factors, such as the type of health facility (CSB1 or CSB2) and whether user fees had been removed (binary variable). In addition, we adjusted for time-varying trends, such as the annual increase in the number of referrals, seasonal trends (using a sinus function with a period of 12 months), and the lagged number of referrals for each health center (lag of one month).

We fit a negative binomial mixed-effects model with all the above variables as fixed effects, and each health center as a random intercept. Model selection was carried out via step-wise selection; where by a multivariate model was first considered including all variables with a p-value below 0.1 in univariate analyses. Then, a reduced model was obtained by keeping only those variables significantly associated with the response variable (i.e., p-values below 0.05). All statistical analyses were carried out using the ‘lme4’ package in R statistical software, version 3.6.2.(41)

#### Predictions of estimated impact for strategies aimed at reducing referral time to secondary care

Given geographic challenges to transport patients from health centers in the North and the South of the district to the district hospital, plans are in place to equip one health center in each area with the necessary infrastructure, equipment, training and human resources to become referral centers. To assess the impact of this strategy, we first estimated health center referral times and household pre-hospital times under a scenario in which health centers refer patients to the closest referral facility among these two new referral centers and the district hospital. Then, using the statistical model of the number of monthly referrals by health center described above, we predicted how reductions in referral time would affect the number of referrals completed in the future. For this, we predicted the number of referrals for a period of two years from the end of the study (July 2020 to June 2022) from the fixed effects of the model using referral times for a scenario with and without the two new centers, and we estimated the difference in the number of referrals between the two scenarios for health centers according to geographic accessibility.

## Results

### Distribution and local determinants of referral and pre-hospital times in Ifanadiana District

Multivariate analyses of over 70,000 km of motorized trips by NGO vehicles in Ifanadiana District revealed that motorized vehicles in the district drove at about 60 km/hon the paved national road (table 1). Speeds decreased by over half on non-paved roads (tertiary or unclassified), and motorcycles drove slightly faster than cars. Travel speed was nearly one-third slower when crossing a bridge or through a residential area. In addition, travel speed was reduced in driving conditions under rain (less than half for every change in a unit of the log_10_of average monthly rainfall) or on a slope (one-third slower for every change in a unit of the log_10_ of % of absolute slope change).

**Table 1.** Multivariate analysis of local factors affecting travel speed by motorized vehicles (Poisson linear mixed model, with individual track as random intercept)

|  | Estimate | 95% Confidence Interval |
| --- | --- | --- |
| **Intercept (km/h)** | 59.98 | [58.15, 61.86] |
| **Type of road** |  |  |
| National road (paved) | (ref) |  |
| Tertiary road (non-paved) | 0.45 | [0.45, 0.45] |
| Unclassified (non-paved) | 0.46 | [0.45, 0.46] |
| **Type of vehicle** |  |  |
| Car/Ambulance | (ref) |  |
| Motorcycle | 1.15 | [1.13, 1.17] |
| **Residential area** |  |  |
| No | (ref) |  |
| Yes | 0.67 | [0.67, 0.67] |
| **Bridge** |  |  |
| No | (ref) |  |
| Yes | 0.70 | [0.69, 0.7] |
| **Distance since trip started (km)** |  |  |
| Within 2km | (ref) |  |
| Over 2 km | 1.18 | [1.18, 1.19] |
| **Slope (% , $\log_{10}$)** | 0.68 | [0.68, 0.68] |
| **Rainfall (mm, $\log_{10}$)** | 0.49 | [0.47, 0.5] |

<sup>1</sup> Model coefficients and confidence intervals are exponentiated to reflect incidence rate ratio (relative change)

Predictions of travel time from this model revealed that only the four health centers located near the paved road had referral times to the hospital within approximately one hour under both dry and rainy conditions (table 2). Two additional health centers reachable by non-paved roads had referral times of one hour under dry conditions, but these increased to about 1h 30min under rainy conditions. The 14 health centers with low accessibility in the North and South of the District required a motorcycle ride or other means of transportation for 20-100 km, plus an ambulance ride for the remaining 17 km to the hospital. Minimum referral time for these health centers varied from 1h 43min to over 6 hours under dry conditions and from 2h 23min to over 8 hours under rainy conditions. These context-specific, data driven estimates of referral time are significantly higher than when using common values of travel speed (50km/h on paved roads and 30km/h on non-paved roads). For instance, referral times for low accessibility health centers were underestimated by a minimum of 30 minutes and a maximum of 1h 45min as compared with our model (table 2).

**Table 2.** Predictions of minimum referral time from each health center to the district hospital under different climatic conditions and predictions with a simple assumption of 50km/h on paved roads and 30km/h on non-paved roads

| Health centers | Accessibility | Distance (km) | Referral time |  |  |
| --- | --- | --- | --- | --- | --- |
|  |  |  | Dry conditions | Rainy conditions | Simplified assumption |
| CSB2 Antaretra | High | 28 (vehicle) | 48min | 1h 6min | 34min |
| CSB2 Ifanadiana |  | 2 (vehicle) | 5min | 7min | 2min |
| CSB2 Kelilalina |  | 9 (vehicle) | 15min | 20min | 11min |
| CSB2 Ranomafana |  | 22 (vehicle) | 33min | 46min | 26min |
| CSB2 Ambiabé | Medium | 14 (vehicle) | 1h 7min | 1h 33min | 28min |
| CSB2 Androrangavola |  | 17 (vehicle), 23 (motorcycle) | 1h 43min | 2h 23min | 1h 6min |
| CSB2 Tsaratanana |  | 17 (vehicle) | 57min | 1h 20min | 34min |
| CSB2 Ambohimanga du Sud | Low CSB2 | 17 (vehicle), 50 (motorcycle) | 3h 15min | 4h 31min | 2h 13min |
| CSB2 Ambohimiera |  | 17 (vehicle), 28 (motorcycle) | 2h 14min | 3h 6min | 1h 29min |
| CSB2 Ampasinambo |  | 17 (vehicle), 110 (motorcycle) | 5h 59min | 8h 19min | 4h 13min |
| CSB2 Analampasina |  | 17 (vehicle), 59 (motorcycle) | 3h 41min | 5h 6min | 2h 31min |
| CSB2 Atsindra |  | 17 (vehicle), 38 (motorcycle) | 2h 42min | 3h 45min | 1h 49min |
| CSB2 Fasintsara |  | 17 (vehicle), 94 (motorcycle) | 5h 15min | 7h 17min | 3h 41min |
| CSB2 Maroharatra |  | 17 (vehicle), 105 (motorcycle) | 5h 45min | 7h 59min | 4h 3min |
| CSB2 Marotoko |  | 17 (vehicle), 31 (motorcycle) | 2h 28min | 3h 25min | 1h 34min |
| CSB1 Ambalavolo | Low CSB1 | 17 (vehicle), 114 (motorcycle) | 6h 14min | 8h 39min | 4h 21min |
| CSB1 AmbodiaraSud |  | 17 (vehicle), 103 (motorcycle) | 5h 43min | 7h 55min | 3h 59min |
| CSB1 Ambodimanga Nord |  | 17 (vehicle), 67 (motorcycle) | 4h 7min | 5h 43min | 2h 47min |
| CSB1 Analamarina |  | 17 (vehicle), 20 (motorcycle) | 1h 52min | 2h 35min | 1h 13min |
| CSB1 Mahasoà |  | 17 (vehicle), 29 (motorcycle) | 2h 3min | 2h 50min | 1h 18min |
| CSB1 Maromanana |  | 17 (vehicle), 52 (motorcycle) | 3h 22min | 4h 40min | 2h 17min |

Combining estimates of referral time by motorized vehicles with previous estimates of walking time from each household to the nearest health center allowed us to estimate the minimum time necessary to seek care at the hospital, or pre-hospital time, at the population level. We found that minimum pre-hospital time varied from 5 minutes to 10 hours under dry conditions and from 8 minutes to 14 hours under rainy conditions. Only 13% of the population lived less than two hours from the hospital and 48% lived within four hours (table 3). This percentage decreased further under rainy conditions, where only 9% and 33% of the population can reach the hospital within two and four hours, respectively. More than half of the population needs over 4h to reach the hospital. As with referral time, comparison of these estimates with commonly used methods for geographic accessibility modeling (friction surfaces, AccessMod) revealed a substantial overestimation of the population with appropriate access to the hospital. For instance, friction surface estimates suggested that nearly 30% of the population could reach the hospital within two hours under dry conditions, and 21.4% under rainy conditions.

**Table 3.** Distribution of the population in Ifanadiana according to their time to reach the district hospital (pre-hospital time)

| Pre-hospital time (hours) | Population (%) |  |  |  |
| --- | --- | --- | --- | --- |
|  | Estimation using statistical model |  | Estimation using AccessMod |  |
|  | Dry conditions | Rainy conditions | Dry conditions | Rainy conditions |
| [0,2] | 12.95 | 9.30 | 29.28 | 21.40 |
| (2,4] | 35.39 | 23.74 | 26.88 | 25.34 |
| (4,6] | 31.13 | 29.28 | 23.26 | 19.82 |
| (6,8] | 14.65 | 19.68 | 8.92 | 15.03 |
| > 8 | 5.88 | 18 | 11.65 | 18.40 |

#### Impact of geographic accessibility on the number of hospital referrals

A total of 8,464 hospital referrals were completed from January 2014 to July 2020. Referral patterns were driven by health center accessibility, with over three quarters of patients referred from the four health centers with high accessibility, and less than 10% of patients referred from the 14 health centers with low accessibility (figure 2, supplementary table S4). The most common means of transportation was by ambulance (40.83%), followed by *taxi brousse* (20.14%) and walking (18.93%). However, the means of transportation varied significantly according to health center accessibility. In health centers with high and medium accessibility more than two thirds of referrals were by ambulance or *taxi brousse*, referrals from health centers with low accessibility required other means (figure 2). About one quarter of referrals from low accessibility facilities required the patient to travel tens of kilometers to a point 17km from the hospital for pickup by the ambulance, another quarter of referrals were completed by foot, and many were completed by tractor or rolling stretchers (figure 2).

**Figure 2.**
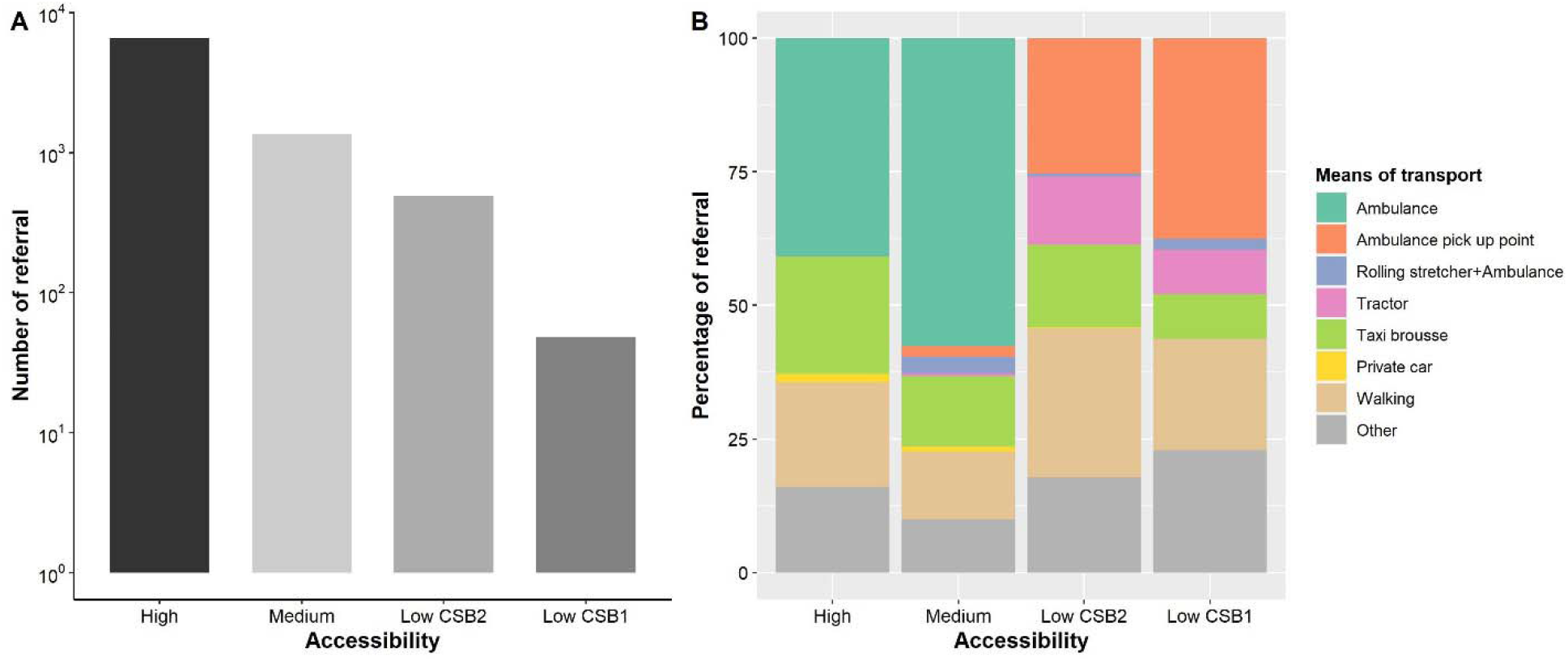
Patterns observed for hospital referrals in Ifanadiana according to geographic accessibility of health centers, 2014-2020. A) Total number of referrals completed during the study period for each category of health center accessibility (y-axis in logarithmic scale) B) Means of referral transport used for each category of health center accessibility.

The number of referrals increased more than three-fold during the study period, from an average of 34 referrals per month in 2014 to 116 referrals per month in the first seven months of 2020 (figure 3). Referral time to the hospital remained the most important determinant of the monthly number of referrals completed by health centers, even after adjusting for other relevant variables (supplementary table S4). We observed an exponential decrease in the number of patients referred as a function of referral time, especially within the first two hours (figure 3). Other factors that were statistically associated with an increase in the number of referrals were the size of the health center (major vs. basic health center), the removal of user fees, and time varying trends (annual increase and lagged number of referrals).

**Figure 3.**
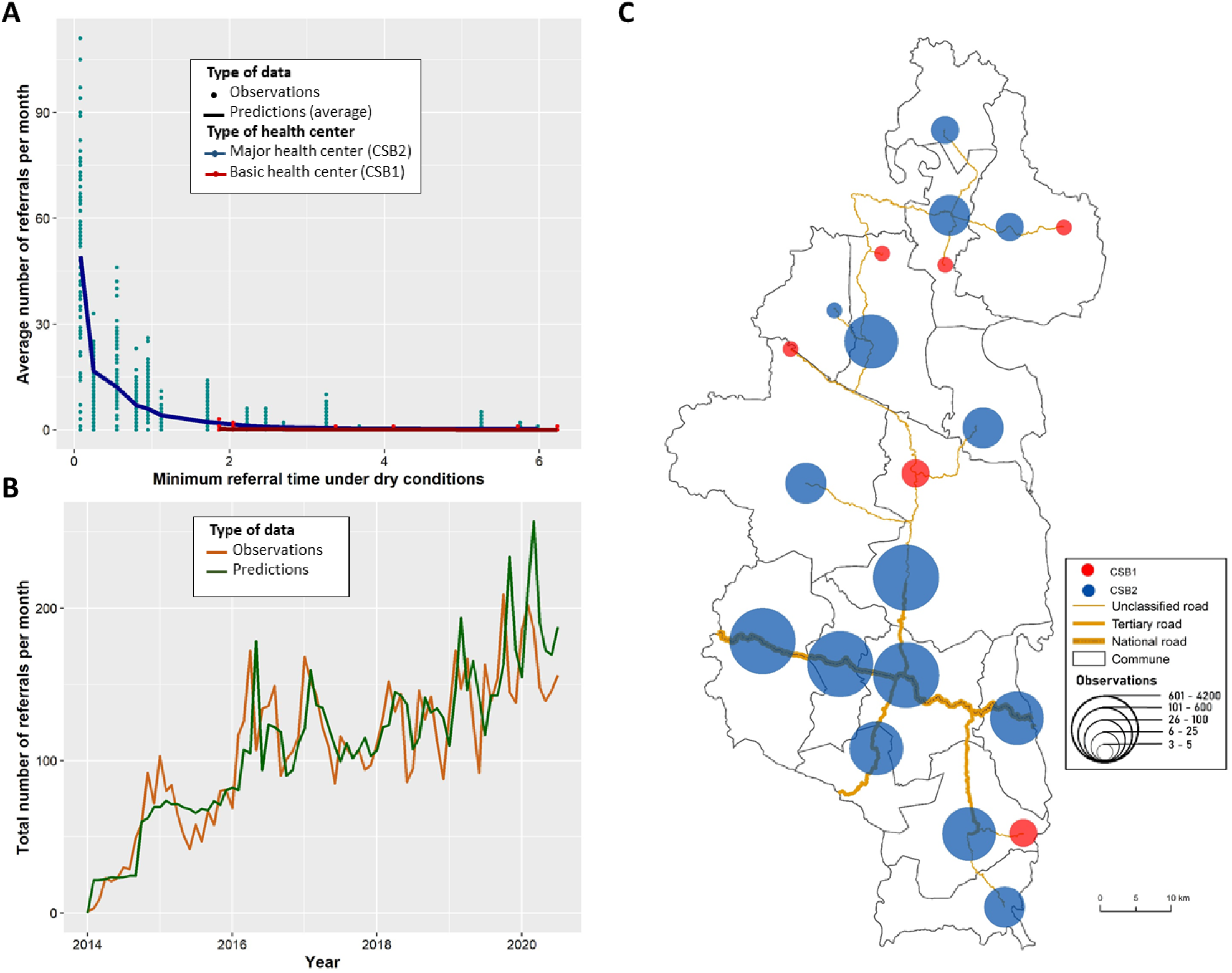
Observed and predicted temporal and spatial trends for hospital referrals in Ifanadiana District, 2014-2020. A) Average number of referrals done by health centers each month as a function of the minimum referral time from the health center to the hospital, stratified by type of health center. B) Evolution of the total number of referrals per month observed (brown line) and predicted (green line) over the study period. C) Spatial distribution of hospital referrals by health center (blue circles= CSB2 and red circles= CSB1), with larger circle size corresponding to a higher number of referrals completed by the health center.

#### Potential impact of strategies that reduce referral time to secondary care

Predictions using locally-calibrated models described above suggested that the implementation of two additional referral centers would lead to a significant increase in access to secondary care, both in terms of a reduction of pre-hospital time and an increase in the number of referrals completed (figures 4 and 5). The main improvements in terms of pre-hospital time would be for the whole Southern and Northern part of the district, where most populations could access a referral center within four hours regardless of climatic conditions (figure 4). This is in contrast to the current situation, in which large geographical areas, especially in the North, have pre-hospital times of over 6hours under dry conditions and over 8hours under rainy conditions (figure 4). The proportion of the population within two hours of a center with higher acute care capacity would increase by nearly three-fold, from 12.95% to 30.56% under dry conditions and from 9.30% to 23.19% under rainy conditions (figure 4). The proportion of the population needing over four hours to reach a health facility would more than halve, decreasing from 51.66% to 24.77% under dry conditions and from 66.96% to 38.16% under rainy conditions.

**Figure 4.**
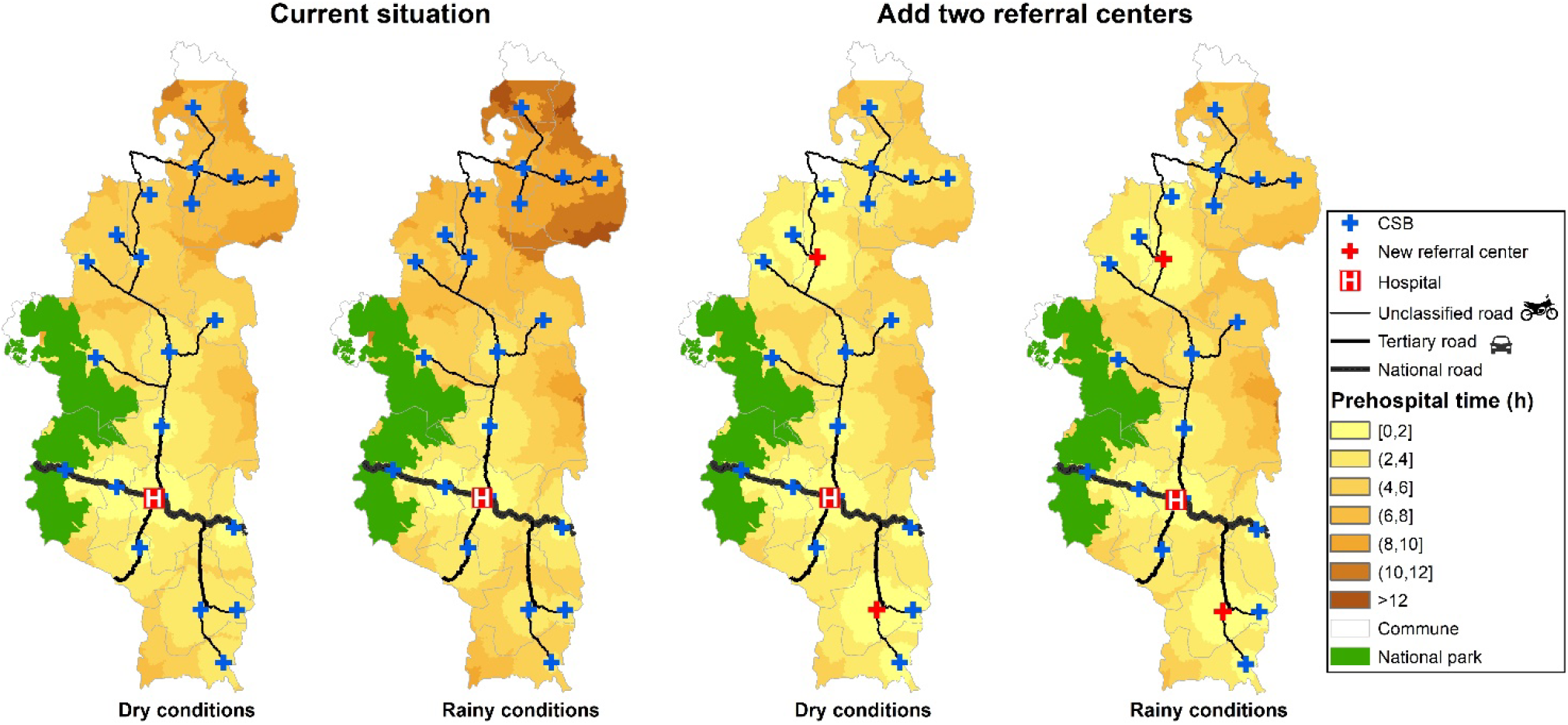
Distribution of the minimum pre-hospital time in Ifanadiana District under different scenarios. Each map shows the time from each of the 41,426 households and residential areas in Ifanadiana to the district hospital (current situation) and under a scenario where two additional referral centersare added (red crosses). Estimates are provided under dry conditions (without rainfall) and rainy conditions (with rainfall). Shading from yellow to brown represent categories of increasing pre-hospital time.

Predictions of the number of referrals for the period from July 2020 to June 2022 suggested that this strategy would lead to a two-fold increase in the number of referrals, from an average of 1906 referrals per year predicted in a scenario with only the district hospital to 3666 referrals per year in a scenario with two new referral centers (figure 5). This increase would disproportionately benefit less accessible health centers over high accessible health centers. While the number of referrals would remain unchanged for health centers with high accessibility at about 30 referrals per month, those with medium accessibility would increase from 8 to 24 referrals per month. For health centers with low accessibility, major health centers (CSB2) would increase monthly referrals from 1 to 13 and basic health centers (CSB1) would increase referrals from 0 to 1 per month.

**Figure 5.**
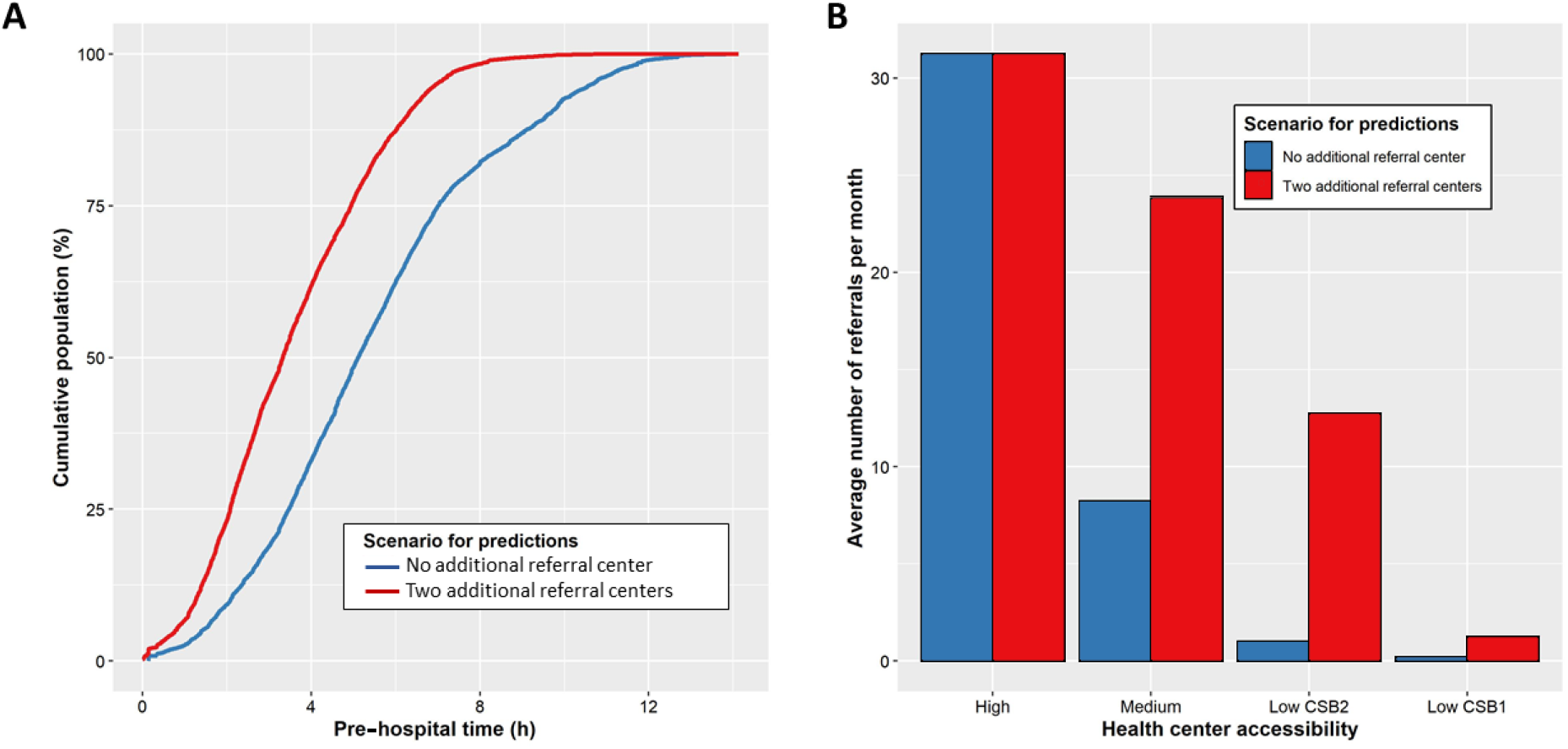
Potential improvements in accessibility to secondary care by adding two referral centers. A Changes in population-level pre-hospital time in a scenario with only one referral hospital (blue) and the addition of two referral centers (red), under rainy conditions B) Changes in the average number of referrals completed by health centers with different levels of geographic accessibility.

## Discussion

In order to address the burden of mortality and disability in LMICs, ensuring access to emergency and hospital care for all has become a global priority, as part of a broader movement to achieve UHC. But many health systems are not designed to reach everyone. As the time necessary to reach a hospital is one of the main determinants to accessing care, it is important to have an accurate understanding of who is able to access service. In this study we combine GPS data of people and motorized vehicles, a full district mapping and remote sensing analyses to calibrate statistical models of local referral and pre-hospital time in a rural district of Madagascar. We showed that only about 10% of the population lives within two hours of the district hospital and more than half live over four hours away, with variable access depending on climatic conditions. Despite health system strengthening efforts in place that have more than tripled the number of hospital referrals between 2014 and 2020, we found that geographic barriers leading to increased referral times remained the primary barrier limiting the number of referrals completed by health centers.

Estimates of geographic accessibility to hospital care for Ifanadiana District were significantly lower than those reported in previous studies. For instance, multi-country studies using friction surface methods combined with available geographic information system (GIS) data had estimated that over 70%(24) and as much as 92%(25) of the population in sub-Saharan Africa lives within two hours of a hospital. Importantly, the scale of these analyses requires trade-offs in the resolution of available input data (e.g. resampling at 1km x 1km) and in the assumptions used (e.g. travel speed for routes and terrains) that likely impact the precision of their travel time estimates. We show here that using simplified assumptions of travel speed for estimating referral time or methods relying on friction surfaces for estimating pre-hospital time in our district led to a substantial underestimation of the travel time necessary to reach the hospital and to an overestimation of the population living within two hours of the hospital. If these local results are representative of larger trends for sub-Saharan Africa, it suggests that results from multi-country studies should be taken with caution. This has major implications for international health policies attempting to reach the recommended goal of ensuring that 80% of the population can access a hospital within two hours, since this threshold may be harder to reach than previously anticipated.

Our study is consistent with previous evidence showing that geographic accessibility to hospital care has a very important role in use.(42,43) Moreover, it reveals that geographic accessibility remains a key barrier even where health system strengthening efforts exist to improve access to hospital care. The MoPH-PIVOT partnership in Ifanadiana included a rare 24/7 ambulance referral system, processes to ensure care continuity such as an emergency call center, and the removal of user fees for referred patients. Yet, over three quarters of hospital referrals in the district came from the four health centers (out of 21) located near the only paved road in the district and within one hour of the hospital. This is consistent with a similar study in Tanzania, where 85.6% of referrals came from highly accessible primary health care centers located within one hour of the hospital.(44) These results reinforce the need to reduce the referral time from health centers to hospitals in LMICs as an effective way to increase hospital use, revealing the limits of health system strengthening programs without further collaboration with other sectors outside the health system. Indeed, improving the quality of local roads, which is beyond the scope of health care NGOs or ministries of health, could significantly expand the geographic reach of ambulances and lead to substantial increases in the number of referrals in addition to having other economic benefits.

We used our models to provide more information on the impact of solutions within the capacity of the MoPH-PIVOT partnership. Previous studies have used geographic accessibility modeling to assess the optimal location of new facilities in order to minimize population travel time.(20,45,46) For instance, in Ethiopia researchers found that upgrading a few strategically located facilities and reconfiguring referral networks could increase the percentage of population served within two hours from 70 to 90%, and reduce the average travel time by nearly half.(20) Another study in Kenya suggested that 32 additional facilities would be necessary to reach UHC for maternity health services in one of Kenya’s rural counties.(45) In this study, we characterized both referral time and its relationship with the number of hospital referrals completed by health centers, which allowed us to inform program managers on the potential impact of a strategy they are planning to implement (i.e., adding two referral centers) not just on pre-hospital time at the population level, but also on the number of expected referrals for each health center. We found that this strategy could be highly effective, doubling the number of hospital referrals in Ifanadiana District, with a notable three-fold increase in referrals for health centers of medium geographic accessibility and more than a 10-fold increase in referrals for health centers with low geographic accessibility. In parallel, the proportion of the district population within two hours from a center with surgical capabilities would nearly triple.

This study had several limitations. First, many scenarios can be considered for how local populations reach a hospital. We assumed that people go to the closest health center by foot and are then referred to the hospital by motorized vehicle because that best reflects local practices in Ifanadiana. However, patients could travel to the hospital by foot, which would increase pre-hospital time (significantly in some cases), or they could come directly to the hospital without passing through the primary health center, which would reduce pre-hospital time. Analyses of referral data showed that less than 20% of referred patients arrived at the hospital by foot, while the fee-exemption program (e.g., costs of referrals are covered for patients sent from a health center to the hospital) in combination with the long distances to reach the hospital strongly incentivized patients to consult at primary health centers first. Therefore, these alternative scenarios probably had a limited impact on the results presented here. Second, predictions of the impact of adding two referral centers on the number of referrals in 2020-2022 were based on trends observed during 2014-2020. If other factors affecting referrals, such as road conditions or health system practices, were to change significantly between these two periods, this could bias our predictions. However, we are not aware that any such changes have taken place to date in our study area. Finally, as with any local study, the generalizability of results presented here may be limited to other rural, low-income settings with similar characteristics as Ifanadiana in terms of terrain, road infrastructure and health system capacity. Further research is needed to assess whether similar trends in hospital geographic access and use are observed in other areas of sub-Saharan Africa.

## Conclusion

This study demonstrates a new method to estimate key measures of geographic accessibility to hospital care that are context-specific and highly accurate. Using high resolution GIS data and local fieldwork, these models can be calibrated to better reflect local practices and barriers in accessing hospital care. Comparisons of results with other commonly used methods in geographic accessibility modeling, suggest that estimates for sub-Saharan Africa may systematically overestimate the percentage of the population living within two hours of a hospital. More broadly, we show how using geographic accessibility modeling methods adapted to local scales by improving the precision of travel time estimates and pairing them with health facility utilization data can inform health actors on ways redesign the local health system to overcome existing barriers to care and improve overall population health.

## Data Availability

Data are available on OpenStreetMap (https://www.openstreetmap.org) and on the Shinny app described in this study (http://research.pivot-dashboard.org:3838/)

http://research.pivot-dashboard.org:3838/

## Acknowledgements

We would like to thank all OSM contributors to the cartography of Ifanadiana District, apart from the authors of this article, and in particular the OpenStreetMap Associations of La Réunion and Madagascar for the mapping parties they organized and their individual contributions. We would also like to thank TAG IP for the motorized vehicle data. In addition, we would like to thank Community Health team staff at PIVOT and community health workers for helping us to collect fieldwork data and PIVOT’s Monitoring and Evaluation staff for the referral data.

## Contributions

FAI, MHB, LR, AG conceived and designed the study. MHB, VH, CR, LFC, AG, advised on the analysis. FAI, MR, AG analyzed the data. FAI, VH, CR, MR, AG contributed to the interpretation of the data. FAI, MHB, MR, LR, VH, CR, MR, JC, SR, GC, ATA, AM, MR, RR, KF, AR, BR, BR, LCF, AG drafted the manuscript. All authors read and approved the final manuscript.

## Patient consent

Not required

## Ethics approval

Not Applicate

## Provenance and peer review

Not commissioned, externally peer reviewed

