## Supplementary file for "Geographic barriers to establishing a successful hospital referral system in rural Madagascar"

**Supplementary table S1:** Summary of HSS interventions implemented in Ifanadiana district between 2014 and 2021, classified by building block of HSS* affected. These included the PIVOT intervention at all three levels of care.

| Level of care | Ifanadiana district (PIVOT catchment+RoD) | |
|---|---|---|
| District hospital Total Ifanadiana: 1 | (1) Network of three ambulances for referrals and emergency care; infrastructure renovations, provision of medical and non-medical equipment, including full laboratory capacity; social support for vulnerable patients<br>(2) Staffing of health workers to reach MoH norms; trainings for medical staff<br>(3) Creation of a hospital-based Monitoring and Evaluation team to follow-up progress of activities<br>(4) Supply chain management and reduction of stock-outs (5) Cost of outpatient and inpatient care fully covered for referred patients<br>(6) Creation of a joint PIVOT-MoH executive committee for hospital management and transparency | |
| | **PIVOT catchment** | **Ifanadiana district (PIVOT catchment+RoD)** |
| Health centres Total Ifanadiana: 21<br>PIVOT catchments: 7 | (1) Infrastructure renovations, provision of medical and non-medical equipment; implementation of IMCI and malnutrition protocols for every child under-five<br>(2) Staffing of health centres above MoH norms; frequent trainings for medical staff<br>(3) Joint MoH-PIVOT training and supervision to improve HMIS data quality<br>(4) Supply chain management and reduction of stock-outs<br>(5) Essential medicines and consumables provided free of charge to all patients<br>(6) Close collaboration with district health managers for the planning and implementation of activities | (1) Provision of medical and non-medical equipment<br>(2) Staffing to bring all health centres up to MoH norms; trainings for medical staff<br>(5) Basic package of health services free of charge for children under-five and pregnant women |
| Community health Total Ifanadiana: 195 fokontany<br>PIVOT catchment: 123 fokontany | (1) Construction of 81 community health sites by community, with PIVOT support; implementation of IMCI and malnutrition protocols for every child under-five<br>(2) Training and on-site supervision of community health workers by mobile teams of trained nurses every 2months (six nurses for ~40CHWs)<br>(3) Joint MoH-PIVOT training to improve HMIS data quality<br>(4) Monthly provision of MNCH medicine stocks to CHWs and follow-up of medicine stock use<br>(5) Cost of MNCH medicine stocks fully covered by PIVOT and provided free of charge to patients; financial incentives to CHWs for stock management and attendance to supervisions (~$4 per month); no fees charged to diagnose patients<br>(6) Community engagement and participation | (1) Provision of non-medical equipment and supplies<br>(2) Training every year; monthly performance evaluation at health centres; on-site coaching by technical assistants (one for ~15–35CHWs) every 2months<br>(4) Provision of a free initial stock of products and medicines (subsequent stocks are purchased by CHWs) †<br>(5) CHWs make a profit from a small margin in the sale of medicines (except those in PIVOT catchment who obtain them for free) †; no fees charged to diagnose patients |

*Building blocks of HSS: (1) service delivery; (2) health workforce; (3) health information systems; (4) medicines and supplies; (5) financing; (6) leadership and governance.

†Exceptionally, these two interventions only happened in RoD, since PIVOT substituted the medicine provision and financial incentives to CHWs. CHW, community health worker; HMIS, health management information systems; HSS, health system strengthening; IMCI, integrated management of child illness; MNCH, maternal, newborn and child health; MoH, Ministry of Health; RoD, rest of the district.

**Supplementary table S2**: Description of the motorized vehicle dataset, which includes GPS recordings of all the NGO vehicles every 10 seconds from March 17 to May 5, 2019 and from February 1 to April 4, 2020.

| | Distance (km) | % |
|---|---|---|
| **Slope (%)** | | |
| [0,30] | 77,229 | 99.38 |
| (30,70] | 428 | 0.55 |
| (70,100] | 52 | 0,67 |
| **Rainfall (mm)** | | |
| [0,10] | 60,650 | 78.05 |
| (10,25] | 11,130 | 1.43 |
| (25,50] | 4350 | 5.60 |
| (50,75] | 1579 | 2.03 |
| **Bridge** | | |
| No | 74,427 | 95,65 |
| yes | 3282 | 4,35 |
| **Residential area** | | |
| No | 68,631 | 88.32 |
| Yes | 9078 | 11.68 |
| **Road** | | |
| National road | 66,443 | 85.50 |
| Tertiary road | 7330 | 9.43 |
| Unclassified | 3936 | 5.07 |
| **Transport** | | |
| Vehicle | 61,258 | 78.82 |
| Motorcycle | 16,258 | 20.92 |

**Supplementary table S3**: Description of hospital referrals in Ifanadiana District, 2014-2020.

| variables | n | % |
|---|---|---|
| **Means of transport** | | |
| Ambulance | 3456 | 40.83 |
| Ambulance pick-up point | 169 | 1.99 |
| Tractor | 74 | 0.87 |
| Taxi-brousse | 1705 | 20.14 |
| Private car | 117 | 1.38 |
| Walking | 1602 | 18.93 |
| Rolling stretcher+Ambulance | 51 | 0.60 |
| Other | 1290 | 15.24 |
| **Accessibility** | | |
| High | 6573 | 77.66 |
| Medium | 1349 | 15.94 |
| Low CSB2 | 494 | 5.84 |
| Low CSB1 | 48 | 0.57 |
| **Year** | | |
| 2014 | 411 | 4.86 |
| 2015 | 816 | 9.64 |
| 2016 | 1423 | 16.81 |
| 2017 | 1403 | 16.58 |
| 2018 | 1465 | 17.31 |
| 2019 | 1783 | 21.07 |
| 2020 (January to July) | 1163 | 13.74 |
| **Season** | | |
| High malaria transmission | 4522 | 53.43 |
| Low malaria transmission | 3942 | 46.57 |

**Supplementary table S4**: Multivariate analysis of local factors affecting the number of monthly referrals by health centers, (negative binomial linear mixed model, with health center as random intercept)

| | **Estimate**[1] | **95% Confidence Intervals** |
|---|---|---|
| Intercept | 0.05 | [0.02, 0.11] |
| **Geographic factors**[2] | | |
| Referral time (hours, linear) | 3.29e-24 | [1.14e-30, 9.49e-18] |
| Referral time (hours, quadratic) | 1.33e+08 | [56.09, 3.15e+14] |
| **Health system factors** | | |
| *Type of health center* | | |
| Basic Health Center (CSB1) | (ref) | |
| Major Health Center (CSB2) | 7.17 | [2.99, 17.21] |
| *Removal of user fees* | | |
| No | (ref) | |
| Yes | 1.11 | [1.08, 1.14] |
| **Time varying factors** | | |
| Linear trend (year) | 2.30 | [1.88, 2.81] |
| Lagged referrals (1-month lag) | 1.02 | [1.01, 1.02] |

[1] Model coefficients and confidence intervals are exponentiated to reflect relative change

[2] Orthogonal polynomial of degree 2. As results are hard to interpret, please refer to Figure 3 to observe changes in the number of referrals according to changes in geography, health system factors and over time.
